# The impact of APOE ε4 in Alzheimer’s disease: a meta-analysis of voxel-based morphometry studies

**DOI:** 10.1101/2024.05.10.24307165

**Authors:** Madison Bailey, Zlatomira Georgieva Ilchovska, Akram A. Hosseini, JeYoung Jung

## Abstract

**Background:** Alzheimer’s disease (AD) is the most prevalent form of dementia, exerting substantial personal and societal impacts. The apolipoprotein E (APOE) ε4 allele is a known genetic factor that increases the risk of AD, contributing to more severe brain atrophy and exacerbated symptoms.

**Purpose:** We aim to provide a comprehensive review of the impacts of the APOE ε4 allele on brain atrophy in AD and mild cognitive impairment (MCI) as a transitional stage of AD.

**Methods:** We performed a coordinate-based meta-analysis of voxel-based morphometry (VBM) studies to identify the patterns of grey matter atrophy in APOE ε4 carriers vs. non-carriers. We obtained coordinate-based structural magnetic resonance imaging (MRI) data for 1135 individuals from 12 studies on PubMed and Google Scholar that met our inclusion criteria.

**Results:** We found significant atrophy in the hippocampus and parahippocampus of APOE ε4 carriers compared to non-carriers, especially within the AD and MCI groups, while healthy controls showed no significant atrophy in these regions.

**Conclusion:** Our meta-analysis sheds light on the significant link between the APOE ε4 allele and hippocampal atrophy in both AD and MCI, emphasizing the allele’s critical influence on neurodegeneration, especially in the hippocampus. Our findings contribute to the understanding of the disease’s pathology, potentially facilitating progress in early detection, targeted interventions, and personalized care strategies for individuals with the APOE ε4 allele who are at risk for Alzheimer’s Disease.

## Introduction

Alzheimer’s disease (AD) remains one of the most pressing challenges in healthcare, imposing significant burdens on individuals, families, and societies worldwide (Lanctot et al. 2024). Currently, there are approximately 50 million AD patients worldwide, with projections indicating a doubling every five years, expected to reach 152 million by 2050 (2022). AD, the most prevalent form of dementia, progresses slowly but inevitably leads to substantial neuronal loss, especially in the medial temporal lobe and hippocampus (Pini et al. 2016), manifesting as memory decline, cognitive impairment, and changes in personality and language abilities (Corey-Bloom 2002; Jahn 2013). Although the disease’s causes are complex, involving genetic, environmental, and lifestyle factors (Eid et al. 2019), the ε4 allele of the apolipoprotein E (APOE) gene emerges as the strongest risk factor for AD (Liu et al. 2013). Approximately 60% of AD patients carry an ε4 allele (Corder et al. 1993; Liu et al. 2013). Carrying one ε4 allele raises the risk of AD by 2-3 times, while those with two ε4 alleles face a 15-fold increased risk of developing AD (Farrer et al. 1997). The presence of the APOE ε4 allele not only increases the risk of developing AD but also can influence the age of onset, progression rate, and severity of the disease (Frisoni et al. 1995; Corder et al. 1996; Cosentino et al. 2008). The current study aims to elucidate the impacts of the APOE ε4 allele on brain structure in AD, drawing upon a comprehensive review of neuroimaging studies.

AD is characterised by the amyloid plaques (Li et al. 2022) and neurofibrillary tangles (Goedert et al. 1991), and these result in synaptic loss, neuronal loss and brain atrophy (Pini *et al*. 2016). In particular, the progressive atrophy of the hippocampus and parahippocampal gyrus disrupts this memory network in AD (Rao et al. 2022). This disruption is further compounded by the accumulation of neurofibrillary tangles and amyloid plaques, the primary pathological features of AD, which contribute to neuronal loss and decreased synaptic functionality (Chu 2012; Josephs et al. 2017). Notably, individuals carrying the APOE ε4 allele show heightened concentrations of amyloid-β42 and tau proteins, correlating with accelerated cognitive decline (Benson et al. 2022).

The APOE gene, located on chromosome 19 (Moreno-Grau et al. 2018) is crucial for synthesizing the APOE, which plays a vital role in transport of cholesterol and other lipids across cells in various tissues in the body (Mahley 1988). In the brain, the APOE protein predominantly produced by astrocytes is critical for the maintenance and repair of neurons (Husain et al. 2021). This gene exists in three main isoforms—ε2, ε3, and ε4—each of which has a unique impact on lipid metabolism and neuronal health (Phillips 2014). The ε4 variant is strongly associated with AD risk, playing a role in the disease’s progression by promoting amyloid β build up and impairing the physiology of the medial temporal lobe (DiBattista et al. 2016). The ε4 allele’s association with increased brain amyloid level is evident in both the early (Caselli et al. 2009; Villemagne et al. 2013) and advanced stages of AD (Nelson et al. 2013; Tai et al. 2013). Additionally, this genetic variant is associated with reduced hippocampal volumes (Manning et al. 2014) and more significant memory deficits in AD patients (Wolk et al. 2010).

Advances in neuroimaging, particularly magnetic resonance imaging (MRI) have been indispensable in identifying and quantifying brain atrophy in AD (Dona et al. 2016). Voxel-based morphometry (VBM) is a neuroimaging analysis technique to investigate focal differences in brain volume using the statistical approach of voxel-wise comparison (Ashburner & Friston 2000). Meta-analysis studies of AD using VBM have focused on comparing AD patients to healthy controls, revealing the atrophy in hippocampus and parahippocampal gyrus (Ferreira et al. 2011; Li et al. 2012; Wang et al. 2015). Despite existing studies on AD, there’s a notable gap in research comparing brain atrophy between APOE ε4 carriers and non-carriers.

Our study aims to fill this gap using a coordinate-based meta-analysis (CBMA) to synthesize data across studies, providing a comprehensive understanding of the APOE ε4 allele’s impact on brain structure through various stages of cognitive decline. We compared brain atrophy in APOE ε4 carriers vs. non-carriers across AD, mild cognitive impairment (MCI), and healthy control (HC) groups. By employing activation likelihood estimation (ALE), we analysed data from multiple studies to identify significant patterns of brain atrophy, offering new insights into the relationship between the APOE ε4 allele and AD pathology.

## Materials and Methods

### Literature search, selection criteria, and quality appraisal

We conducted a comprehensive literature search from Oct 2023 to Feb 2024. This search spanned PubMed and Google Scholar, aiming to find structural neuroimaging studies written in English that compare APOE ε4 carriers with non-carriers. Our search strategy involved the following combination of keywords: 1) (Alzheimer) AND (voxel-based morphometry or VBM) AND APOE (apolipoprotein or APOE), 2) (mild cognitive impairment or MCI) AND (voxel-based morphometry or VBM) AND APOE (apolipoprotein or APOE).

This search strategy yielded 83 studies, of which 12 were relevant to the following inclusion criteria: i) studies that used VBM as the imaging modality; ii) studies that reported brain coordinates in a standardised stochastic space, either Montreal Neurological Institute (MNI) or Talairach templates; iii) studies that compared group contrasts between APOE ε4 carriers and non-carriers; iv) studies that reported atrophy of grey matter; v) studies that performed a whole-brain analysis. After the initial screening process, 33 studies were eligible for assessment; of these, we excluded 24 for the following reasons: i) comparative studies (n = 6); ii) reviews (n = 1); iii) case reports (n = 1); iv) no genetic information (n = 4); v) no contrast information (n = 2); vi) white matter studies (n = 2); vii) no whole brain analysis (n = 2); viii) non-human studies (n = 3). Figure 1 illustrates how we applied the inclusion and exclusion criteria to the study selection process using the PRISMA (Preferred Reporting Items for Systematic Reviews and Meta-Analyses) 2020 flow diagram for new systematic reviews {Page, 2021 #105}. MB, ZI, and JJ conducted all steps. The articles that met the criteria after the screening process were included in the meta-analysis.

**Figure 1.**
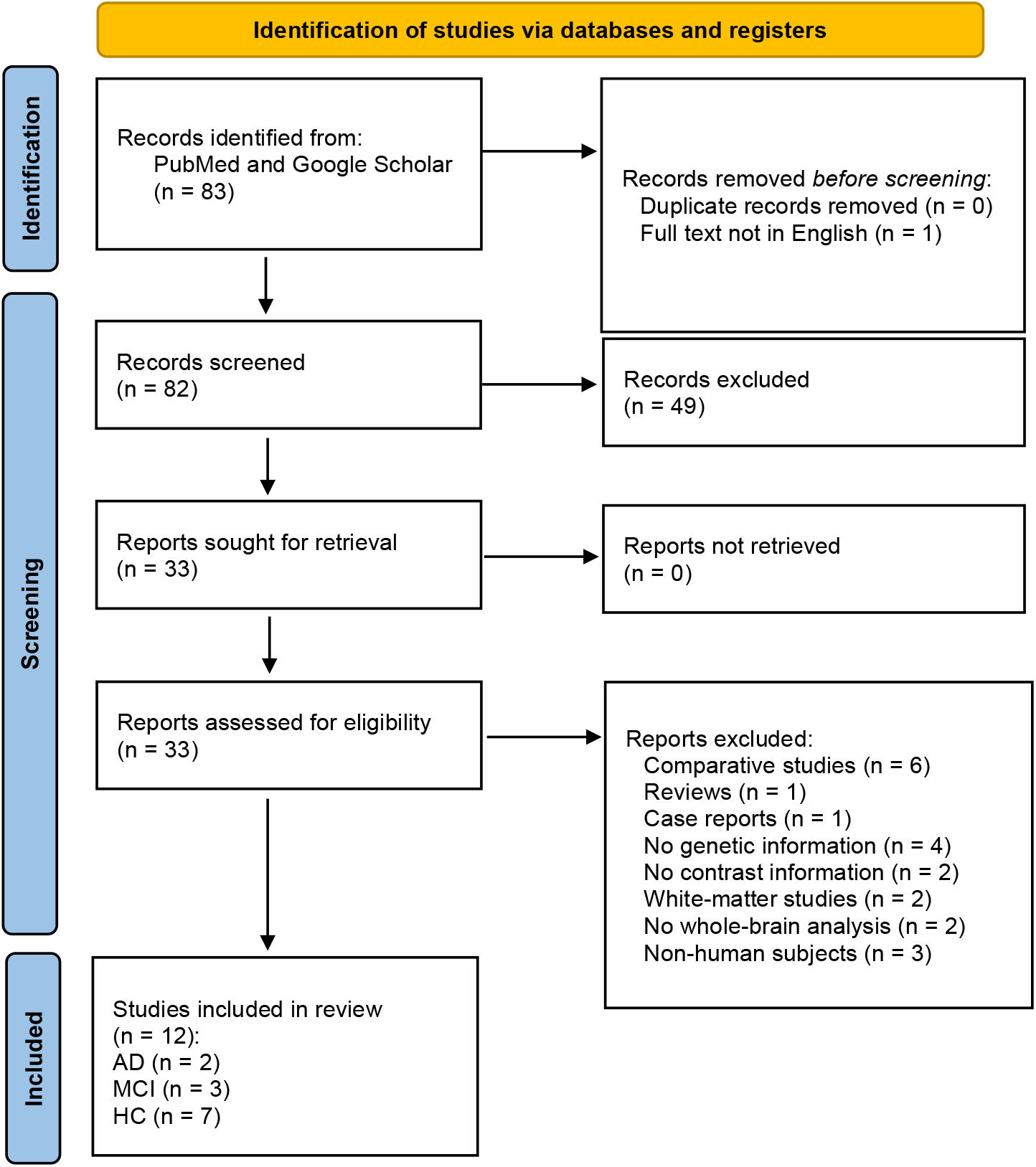
PRISMA flowchart of study selection.

### Activation likelihood estimation (ALE) analysis

For a comprehensive investigation into the impact of APOE ε4 status on brain atrophy, we conducted meta-analysis using ALE algorithm by BrainMap GingerALE 3.0.2 (https://brainmap.org) {Eickhoff, 2009 #78}. The experiments were categorised into AD, MCI, and HC groups (Table 1). Separate ALE meta-analyses were performed on four subsets of the experiments: (1) all patient experiments combined (AD + MCI); (2) experiments per each group (AD, MCI, and HC). Some studies included multiple experiments; hence, the total number of experiments (n = 25) exceeds the total number of studies (n = 12). All Talairach coordinates were transformed into MNI space. Threshold was set to uncorrected p < 0.001 with a 1000 permutation. The minimum cluster volume was set to 200 mm^3^.

**Table 1.**
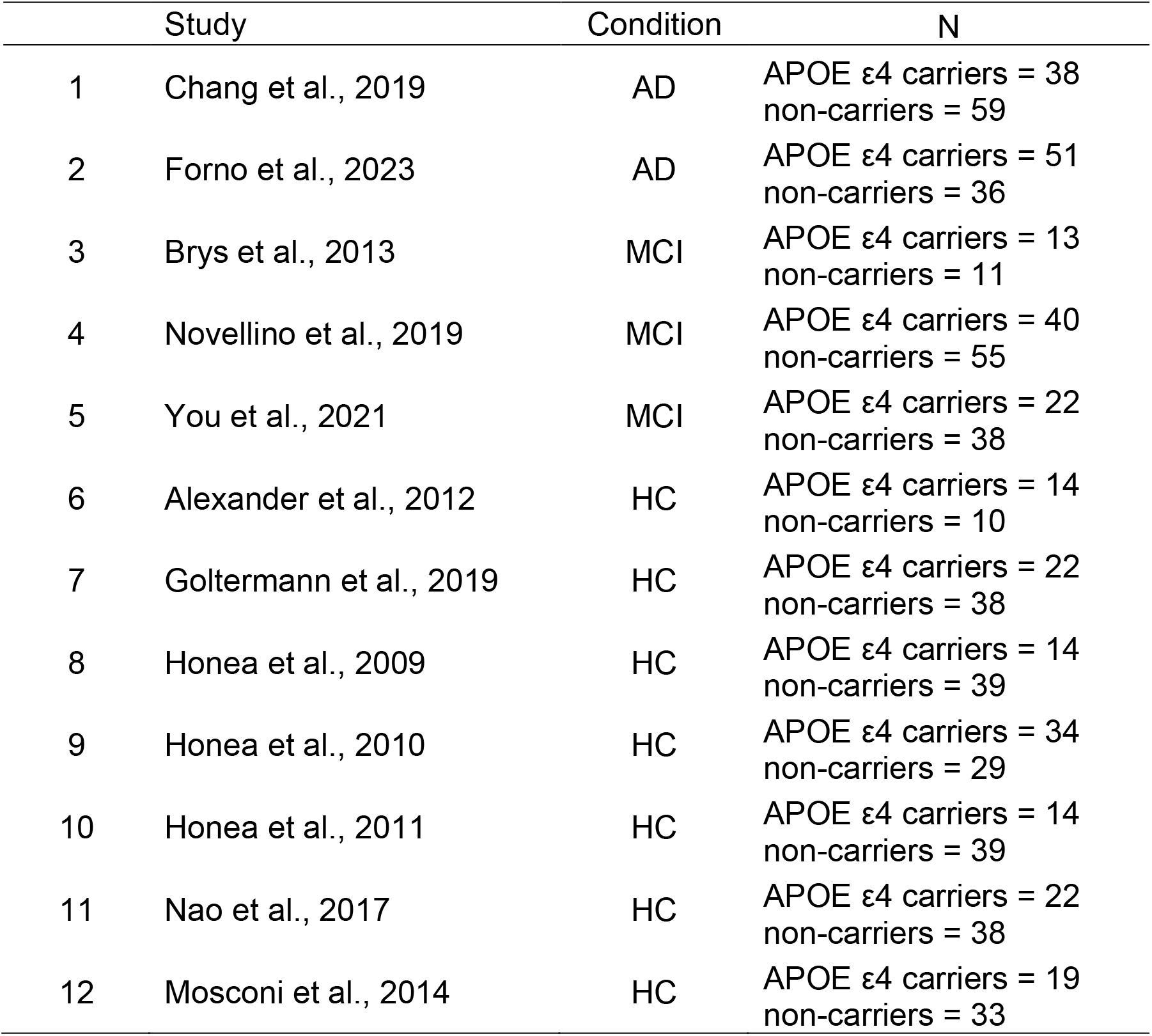
The list of 12 studies included in the meta-analysis.

### Neurosynth

To link the patterns of brain atrophy identified from our meta-analysis to functional networks, we performed additional meta-analyses using Neurosynth (Yarkoni et al. 2011) (http://neurosynth.org). We selected the peak coordinates from each cluster identified in our meta-analysis as a seed region to create meta-analytic coactivation maps and to identify keywords related to these maps. Neurosynth generates a z-score map, indicating the probability that a specific term was mentioned in a research article in association with an activation site. We applied a voxel-wise threshold to the Neurosynth outcomes at a significance level of 0.01, adjusted for false discovery rate (FDR). Keywords were selected based on their z-scores being greater than 0, with up to 5 keywords chosen according to their z-score magnitude.

## Results

### ALE meta-analysis results

Twelve studies, 25 experiments with 1135 participants, yielding 164 foci met the relevant inclusion criteria and were entered into the analysis. The results are summarised in Fig. 2 and Table 2. The primary analysis with all patients (AD + MCI, 10 experiments, 46 foci, 608 patients) revealed that patients carrying the APOE ε4 allele exhibited significant atrophy in the bilateral hippocampus and paraphippocampal regions compared to those without the allele (Fig. 2A). Further detailed analyses by patient category revealed that, in AD patients (6 experiments, 35 foci, 405 ADs), APOE ε4 carriers showed notable atrophy in the bilateral hippocampus, the right parahippocampal gyrus, and the posterior cingulate cortex (Fig. 2B). In MCI patients (4 experiments, 11 foci, 203 MCIs), carriers of the APOE ε4 allele exhibited atrophy in the bilateral paraphippocampal gyrus and the globus pallidus (Fig. 2C). Among HCs (12 experiments, 113 foci, 527 HCs), those with the APOE ε4 allele had atrophy in the right superior temporal gyrus (STG) (Fig. 2D).

**Figure 2.**
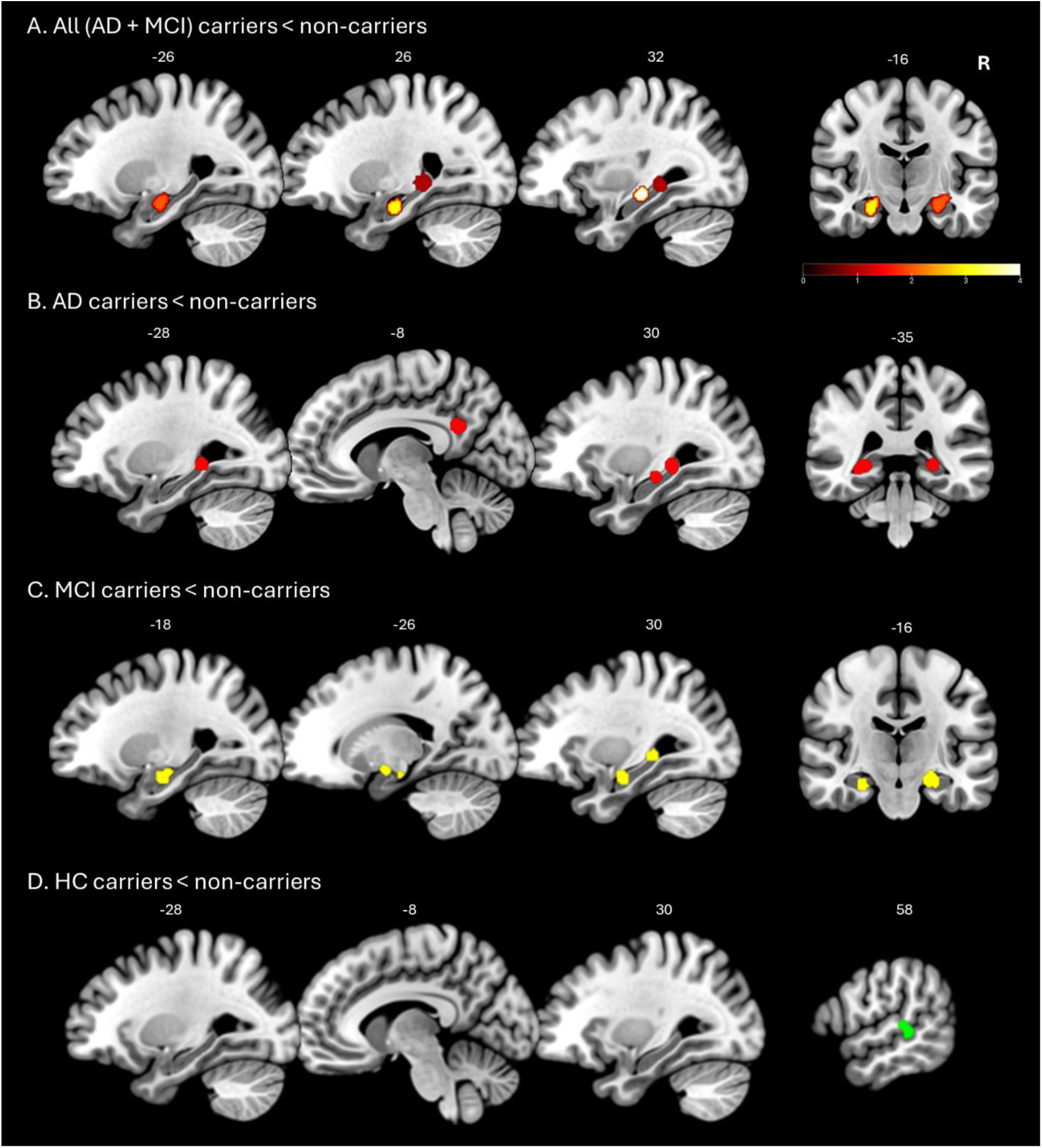
The results of ALE meta-analysis. A) The results of all patients (AD + MCI) carriers < non-carriers. B) The results of AD carriers < non-carriers. C) The results of MCI carriers < non-carriers. D) The results of HC carriers < non-carriers. Colour bar indicates Z-score.

**Table 2.**
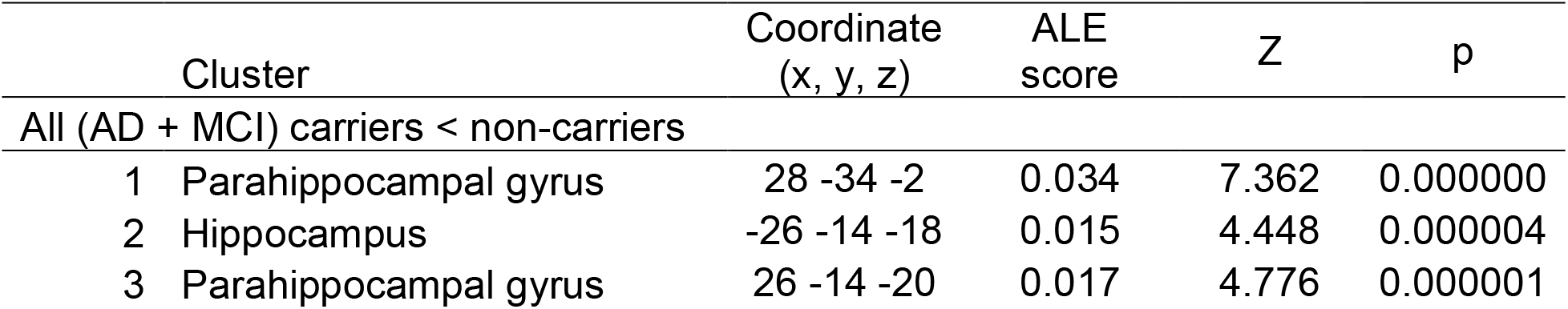
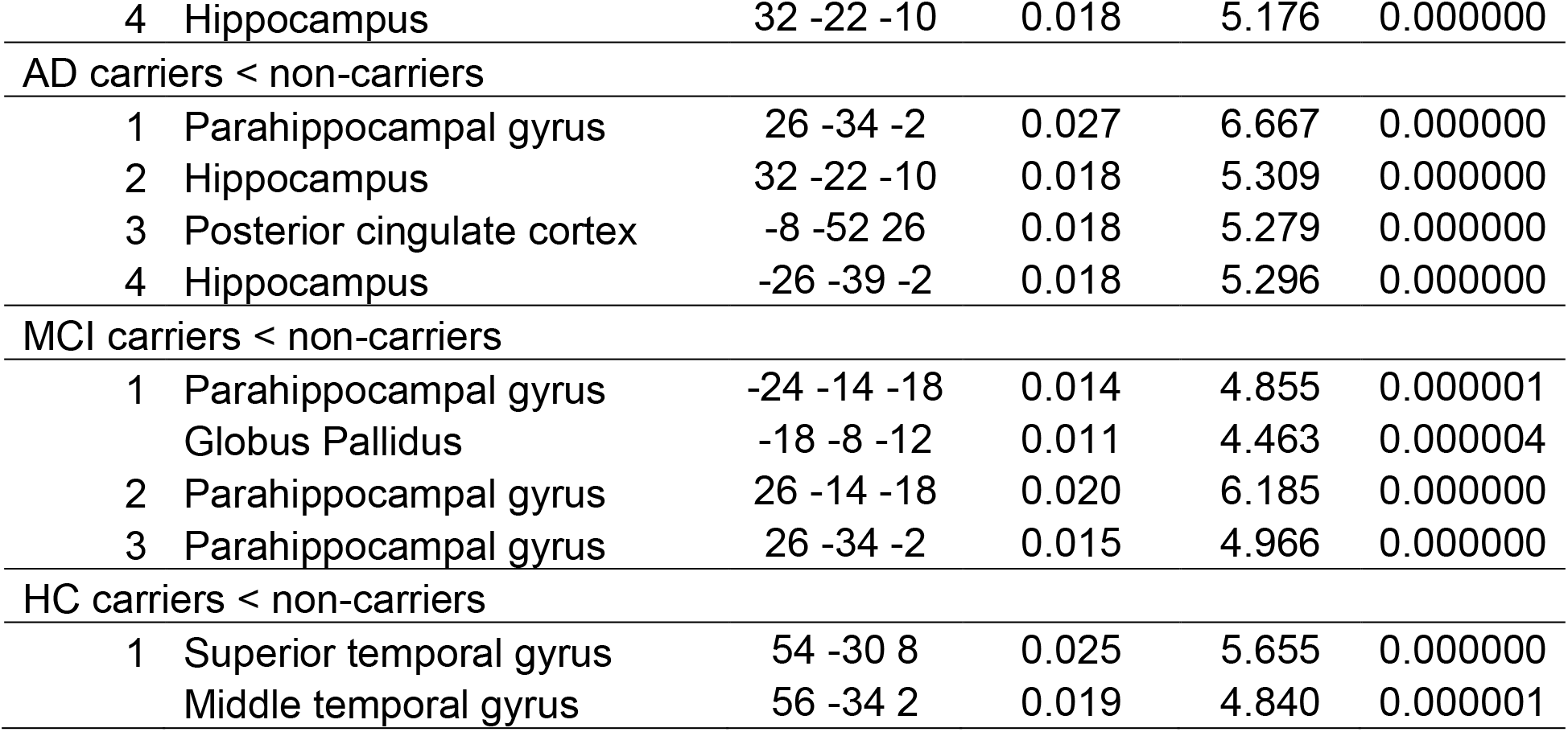
The results of ALE meta-analysis.

| Cluster | Coordinate (x, y, z) | ALE score | Z | p |
| --- | --- | --- | --- | --- |
| All (AD + MCI) carriers < non-carriers |  |  |  |  |
| 1 Parahippocampal gyrus | 28 -34 -2 | 0.034 | 7.362 | 0.000000 |
| 2 Hippocampus | -26 -14 -18 | 0.015 | 4.448 | 0.000004 |
| 3 Parahippocampal gyrus | 26 -14 -20 | 0.017 | 4.776 | 0.000001 |

Table 2. The results of ALE meta-analysis
|  |  |  |  |  |  |
| --- | --- | --- | --- | --- | --- |
| 4 | Hippocampus | 32 -22 -10 | 0.018 | 5.176 | 0.000000 |
| AD carriers < non-carriers |  |  |  |  |  |
| 1 | Parahippocampal gyrus | 26 -34 -2 | 0.027 | 6.667 | 0.000000 |
| 2 | Hippocampus | 32 -22 -10 | 0.018 | 5.309 | 0.000000 |
| 3 | Posterior cingulate cortex | -8 -52 26 | 0.018 | 5.279 | 0.000000 |
| 4 | Hippocampus | -26 -39 -2 | 0.018 | 5.296 | 0.000000 |
| MCI carriers < non-carriers |  |  |  |  |  |
| 1 | Parahippocampal gyrus | -24 -14 -18 | 0.014 | 4.855 | 0.000001 |
|  | Globus Pallidus | -18 -8 -12 | 0.011 | 4.463 | 0.000004 |
| 2 | Parahippocampal gyrus | 26 -14 -18 | 0.020 | 6.185 | 0.000000 |
| 3 | Parahippocampal gyrus | 26 -34 -2 | 0.015 | 4.966 | 0.000000 |
| HC carriers < non-carriers |  |  |  |  |  |
| 1 | Superior temporal gyrus | 54 -30 8 | 0.025 | 5.655 | 0.000000 |
|  | Middle temporal gyrus | 56 -34 2 | 0.019 | 4.840 | 0.000001 |

### NeuroSynth results

To explore the functional networks linked with the patterns observed in our meta-analysis, specifically comparing AD carriers to non-carriers, we utilized Neurosynth for a meta-analytic coactivation analysis. The results are summarized in the Fig. 3. Analysis of the coactivation map for the parahippocampal gyrus cluster (MNI coordinates 26, -34, -2) uncovered connectivity with the bilateral hippocampus, parahippocampal gyrus, and posterior cingulate cortex (PCC) (Figure 3A). The hippocampal clusters (MNI 32, -22, -10 and -26, -39, -2) were primarily linked with the bilateral hippocampus (Fig. 3B and C). The coactivation analysis of the posterior cingulate cortex showed its association with the medial prefrontal cortex, precuneus, posterior cingulate cortex, bilateral angular gyrus, middle temporal gyrus, and hippocampus (Fig. 3D). Additionally, word clouds generated from Neurosynth meta-analysis decoding revealed the most frequently associated terms with the brain activity patterns in these coactivation maps, including ‘episodic’, ‘memory’, ‘encoding’, ‘retrieval’, among others (Fig. 3E).

**Figure 3.**
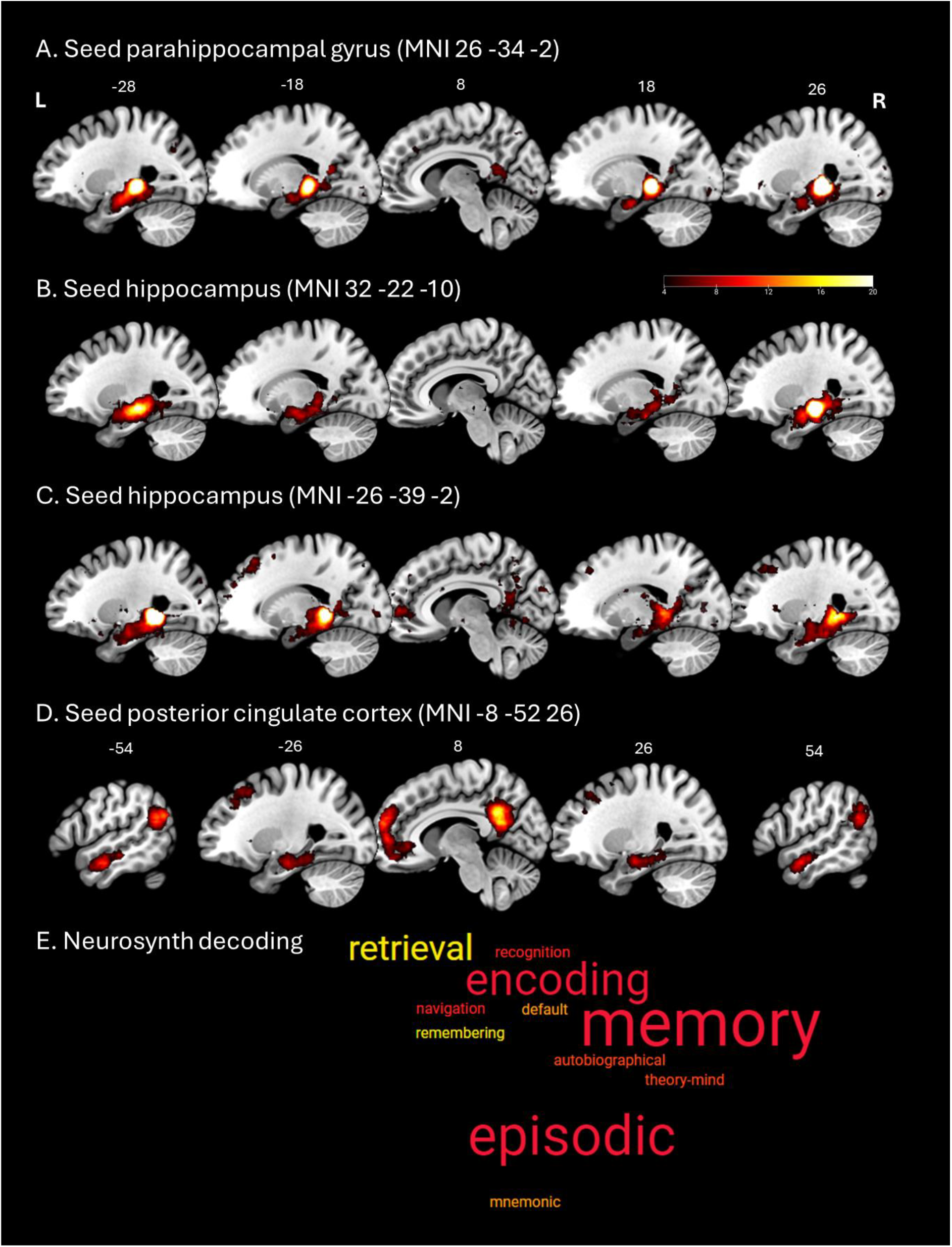
The results of NeuroSynth. A) The meta-analytic coactivation map of the parahippocampal gyrus cluster. B) The meta-analytic coactivation map of the right hippocampus cluster. C) The meta-analytic coactivation map of the left hippocampus cluster. D) B) The meta-analytic coactivation map of the posterior cingulate cortex cluster. E) Word clouds illustrate the findings of a meta-analysis conducted with Neurosynth, showing the terms most commonly associated with the brain activity patterns of seed regions identified when comparing AD carriers to non-carriers. Colour bar represents Z-score.

## Discussion

Our meta-analysis aimed to explore how the APOE ε4 allele influences brain atrophy in AD, offering insights into the underpinning of brain atrophy in this condition. By examining the effects of the APOE ε4 allele in individuals at various stages of clinical progression, AD and MCI, we revealed the APOE ε4 allele’s impact on the hippocampus and parahippocampus, areas vulnerable to neurodegenerative changes. Our findings demonstrated significant atrophy in the hippocampus and parahippocampus of APOE ε4 carriers compared to non-carriers, especially within the AD and MCI groups, while healthy controls despite being carriers showed no significant atrophy in these regions. This finding is consistent with existing research highlighting the hippocampus’s susceptibility to early degenerative changes in AD, where it plays a critical role in memory formation, storage, and retrieval (Ferreira *et al*. 2011; Li *et al*. 2012; Wang *et al*. 2015; Pini *et al*. 2016). The observed atrophy in these critical memory regions among APOE ε4 carriers suggests the allele may contribute to the exacerbation of memory loss in AD, supporting the notion that this genetic variant is a significant risk factor for accelerated neurodegeneration (Corder *et al*. 1993; Liu *et al*. 2013; Lancaster et al. 2019).

The APOE ε4 allele’s presence has been associated with diminished volumes in the hippocampus, particularly evident in older individuals with AD and MCI, revealing an average volume decrease of about 4% in the hippocampus per APOE ε4 allele (Fleisher et al. 2005; Fleisher et al. 2013; Chauhan et al. 2015). Utilizing findings from large-scale Genome Wide Association Study Meta Analyses (GWASMA), Lupton et al (2016) further confirmed the link between the APOE ε4 allele and hippocampal volume reduction in AD and MCI, indicating that carriers of the ε4 allele exhibit smaller hippocampal volumes than non-carriers. Moreover, Kerchner et al (2014) reported that APOE ε4 carriers in MCI and AD patients had thinner hippocampal subregions and showed poorer episodic memory abilities compared to non-carriers. Moreover, our NeuroSynth analysis demonstrated the hippocampal network associated with memory function. Located in the medial temporal lobe, the hippocampus is among the initial regions to exhibit damage in AD (Pini *et al*. 2016). This early vulnerability is attributed to its critical role in encoding new memories and navigating spatial environments (Bird and Burgess 2008). The parahippocampal gyrus, surrounding the hippocampus, supports memory encoding and retrieval by processing environmental contextual information (Aminoff et al. 2013). Together, these structures are crucial components of the brain’s memory network, facilitating the transition of short-term memories to long-term storage (van Strien et al. 2009). Our meta-analysis corroborates these genomic and neuroimaging findings, demonstrating the significant influence of the APOE ε4 allele on hippocampal atrophy within AD and related their memory impairments.

Furthermore, our analysis revealed atrophy in the posterior cingulate cortex (PCC) among AD patients carrying the APOE ε4 allele compared to non-carriers. The PCC is an integral part of the Papez circuit, which also includes the hippocampus, mammillary bodies, thalamus, and parahippocampal gyrus (Aggleton et al. 2022). This circuit plays a pivotal role in memory formation, emotional regulation, and is implicated in the episodic memory and spatial navigation challenges seen in AD (Forno et al. 2021). The PCC is also a central element of the default mode network (DMN), which is known to be affected by AD pathology (Callen et al. 2001; Buckner et al. 2008). Studies repeatedly have reported the decreased volume of PCC (Callen *et al*. 2001) and disrupted functional connectivity of the PCC in AD (Buckner *et al*. 2008). Importantly APOE ε4 carriers, both in AD and MCI stages, have been observed to exhibit loss of grey matter in the PCC (Haller et al. 2017) and impaired functional connectivity (Sheline et al. 2010). Aligned with existing evidence, our findings suggest the influence of the APOE ε4 allele on PCC abnormalities, supporting he notion of PCC atrophy as a distinctive cortical feature of AD (Dickerson et al. 2009). Overall, our results highlight the contribution of APOE ε4 carriers at genetic risk for AD to the brain atrophy and the cognitive deficits associated with AD.

Despite the robust associations observed, our study’s limitations, including a relatively small sample size and the scarcity of studies directly comparing APOE ε4 carriers to non-carriers, necessitate cautious interpretation of our results. This study is not a mechanistic study, and more studies are needed to better understand if the APOE has a direct or indirect impact on the Alzheimer’s pathology, focal brain atrophy/neurodegeneration, or conversion of MCI to AD, or disease progression. However, our analysis lays a foundational basis for future research in this field, suggesting the importance of genetic screening in tailoring potential prevention and treatment strategies for AD. APOE genetic test is also important for monitoring the amyloid-therapy (e.g. Donanemab, Lecanemab), and it could be useful to study the influence of amyloid therapy in slowing down the hippocampal atrophy in the carriers vs non-carriers. Recognizing the multifactorial nature of AD, further studies are essential to unravel the complex interplay of genetic and neuronal factors underlying the disease, providing the way for more effective interventions and a deeper understanding of its pathophysiology.

By performing a meta-analysis on structural neuroimaging data, we showed how this genetic variant contributes to the understanding of disease and its impact on brain structure. By identifying specific brain atrophy patterns associated with the APOE ε4 allele, our meta-analysis provides the significant link between genetic factors and neurodegeneration, offering potential pathways for targeted interventions and treatments. The recognition that APOE ε4 carriers in AD exhibit distinct patterns of brain atrophy, especially in the hippocampus and Papez circuit, enables more accurate risk assessment and individualized approaches to mitigating neurodegenerative progression.

## Data Availability

All data produced in the present work are contained in the manuscript.

## Acknowledgements

This research was supported by AMS Springboard (SBF007\100077) to JJ. AAH has received funding from the Medical Research Council, UK (Grant <u>MR/T005580/1</u>) and National Institute of Health/NIA, USA (Grant <u>1R56AG074467-01</u>). She has received honoraria from Biogen, Eisai, and Lilly for advisory consultations and teaching related to Alzheimer’s disease.

